# Accuracy of Online Symptom-Assessment Applications, Large Language Models, and Laypeople for Self-Triage Decisions: A Systematic Review

**DOI:** 10.1101/2024.09.13.24313657

**Authors:** Marvin Kopka, Niklas von Kalckreuth, Markus A. Feufel

## Abstract

Symptom-Assessment Application (SAAs, e.g., NHS 111 online) that assist medical laypeople in deciding if and where to seek care (*self-triage*) are gaining popularity and their accuracy has been examined in numerous studies. With the public release of Large Language Models (LLMs, e.g., ChatGPT), their use in such decision-making processes is growing as well. However, there is currently no comprehensive evidence synthesis for LLMs, and no review has contextualized the accuracy of SAAs and LLMs relative to the accuracy of their users. Thus, this systematic review evaluates the self-triage accuracy of both SAAs and LLMs and compares them to the accuracy of medical laypeople. A total of 1549 studies were screened, with 19 included in the final analysis. The self-triage accuracy of SAAs was found to be moderate but highly variable (11.5 – 90.0%), while the accuracy of LLMs (57.8 – 76.0%) and laypeople (47.3 – 62.4%) was moderate with low variability. Despite some published recommendations to standardize evaluation methodologies, there remains considerable heterogeneity among studies. The use of SAAs should not be universally recommended or discouraged; rather, their utility should be assessed based on the specific use case and tool under consideration.

## Introduction

Symptom-assessment Applications (SAAs, also known as online symptom checkers or digital triage tools) are digital platforms accessible via smartphones or websites that analyze symptoms using various methods^1,2^. They provide a diagnosis and recommendation whether and where medical care should be sought, a process referred to as *self-triage*^2^. SAAs are potentially useful for various stakeholders: health protection agencies may use the symptom input for syndromic surveillance^3^, general practitioners and clinics can implement SAAs for patient (re-)direction^4,5^ and medical laypeople can use them for assistance in health-related decisions^6^. Hence, they could make healthcare resource distribution more efficient and ultimately increase healthcare access and health equity by providing health advice and recommendations regardless of a person’s socioeconomic status, education, or other determinants of health.

SAAs are increasingly used worldwide. For instance, the United Kingdom’s National Health System (NHS) launched *NHS 111 online* in 2017^7^ and Germany’s Association of Statutory Health Insurance Physicians supplemented their triage hotline with the digital *PatientenNavi* in 2021^8,9^. Consequently, these tools perform millions of assessments annually, with about 7% of the German population using SAAs^7,10^. However, some studies raised concerns about their real-world utility and cost-effectiveness, as they did not seem to reduce healthcare utilization in an NHS evaluation study^11^. This is no surprise, as SAAs tend to be risk-averse and frequently provide users with a recommendation of higher urgency than necessary, making them seek care more often^2,12^. The opposite of this *over-triage* is *under-triage* – where users receive a recommendation of lower urgency than warranted – and poses potential safety risks to users^12,13^. Hence, both the safety and accuracy of SAAs have been subjects of several studies. Three systematic reviews have been published to synthesize the available evidence on SAAs so far and show that SAA accuracy is generally far from perfect, but they demonstrate a high variability between different apps^14–16^.

As an alternative to SAAs, Large Language Models (LLMs) have been proposed in some studies^13,17^. After becoming available in 2022, they quickly garnered interest in the medical community for passing state licensing exams and, as a result, are now suggested as potential clinical decision support system ^18–20^. Some studies have also tested LLMs with cases developed for SAAs and suggest them as decision support tools for medical laypeople as well^21,22^. Nevertheless, an evidence synthesis that reports the accuracy of LLMs for self-triage decisions is still missing.

All these studies on SAAs and LLMs have in common that they view these tools as sole decision-makers, and most researchers recommend or discourage their use without considering the accuracy of actual users. This perspective might overlook scenarios where – if users alone perform poorly – even suboptimal SAAs could be beneficial. Conversely, if users generally make very good decisions, SAAs might not offer any effective assistance. Although one study compared the accuracy of SAAs directly with that of laypeople^23^, an evidence synthesis contextualizing the accuracy of SAAs and LLMs with the accuracy of laypeople is missing.

Therefore, this systematic review aims to extend previous reviews on SAA accuracy^14–16^ by including studies on the accuracy of LLMs as an alternative to SAAs and medical laypeople as users. This comparison shifts the focus from SAAs and LLMs as the sole decision-making entity to considering their user group of medical laypeople as a benchmark against which their accuracy should be interpreted. Since specific diagnoses are of no use for medical laypeople – and are ultimately made and treated by medical professionals anyway – this review focuses on self-triage decisions only and deliberately excludes diagnostic accuracy to focus on user utility.

## Methods

### Eligibility Criteria

This study was preregistered on PROSPERO (ID: CRD42024563111) and adheres to the PRISMA reporting guideline^24^. Following a previous systematic review on SAAs^16^, we included studies published from 2010 onward. We included all primary research articles (including preprints) that were published in English language. Our inclusion criteria comprised all patient demographics (including both vignette-based and real-world evidence studies) and various symptoms, but we excluded studies that focused solely on highly specialized tools or cases, such as only COVID-19 SAAs or COVID-19 cases only^25^. Our inclusion criteria required interventions to examine the self-triage advice of SAAs, LLMs, or laypeople. We excluded any studies that evaluated multiple tools being used simultaneously (e.g., SAAs combined with a telephone triage hotline) or tools that did not offer self-triage advice. Each study needed a gold standard solution for each case as a comparator. Studies that only rated the appropriateness of the received self-triage advice (e.g., on a 5-point Likert scale) without providing a direct solution to a case were excluded. Lastly, studies were required to quantitatively report (self-)triage accuracy by advising the most appropriate care facility, as this recommendation is the purpose that SAAs are developed for^2^. We excluded any studies that exclusively reported triage accuracy for emergency departments (e.g., using the Manchester Triage Scale or Emergency Severity Index) without considering other care facilities. Studies that reported only diagnostic accuracy without corresponding self-triage accuracy were excluded as well.

For the synthesis, we grouped studies according to the agent for which they provided self-triage accuracy estimates, i.e., for SAAs, LLMs and/or laypeople.

### Search Strategy & Information Sources

We conducted our search on July 09, 2024, using the databases Web of Science, MEDLINE / Pubmed, and Scopus to identify relevant articles. The search was limited to studies published from 2010 onward and included English articles only. We developed an initial search string based on previously published systematic reviews of SAAs^14–16^ and adapted it to focus on self-triage accuracy and to include LLMs and laypeople. This search string was refined until it identified all studies reporting self-triage that previous systematic reviews reported. The same refined search string was applied across all databases. The search string for Web of Science read:

> *AB=(app OR apps OR application OR artificial intelligence OR AI OR online OR web-based OR chatbot OR mobile OR computer-assisted OR internet OR smartphone OR phone OR web) OR AB=(symptom checker OR symptom check* OR symptom assessment app* OR symptom-assessment app* OR webmd OR symptomate OR ada OR yourmd OR mediktor OR buoy OR self-refer*) OR AB =(human OR layperson OR laypeople OR lay OR user OR non-professional OR non-clinician) OR AB =(GPT-3 OR ChatGPT OR GPT-4 OR GPT-4o OR Large Language Model OR LLM OR Claude OR Google Bard OR Mistral OR GPT)) AND AB =(self-triage OR triage OR symptom urgency OR dispositional advice OR self-assess*) AND AB=(accuracy OR correct)*

After identifying relevant articles, we conducted both forward and backward citation searches to identify additional studies, particularly preprints, that were not initially retrieved from the databases.

### Data Extraction & Data Analysis

The studies were retrieved and imported into PicoPortal, where they were deduplicated. The titles and abstracts were screened by two researchers (MK & NvK) independently on PicoPortal. In cases of disagreement, both researchers re-examined the title and abstract and resolved conflicts through discussion. Afterwards, the full texts of each eligible study were independently screened by both researchers according to the pre-specified inclusion and exclusion criteria. Cases of disagreement were examined again, and conflicts were resolved through further discussion.

The data were extracted by both researchers independently using a standardized Excel template. The primary outcome focused on assessing the self-triage accuracy of SAAs, LLMs, and laypeople. For the secondary outcomes, the researchers extracted reported accuracy across the urgency levels and the specific self-triage accuracy of each individual SAA and LLM. To gain insights into differences in methodology, the data extraction form included the number of SAAs, LLMs, and laypeople, a brief description of the methods used, the number of triage levels in the study, the number of cases examined, the gold standard assignment process, the number of data inputters, other reported outcomes with respect to self-triage, as well as any conflicts of interest and funding sources. Any instances of missing data were coded as ‘*not available’*. Due to the varying methodologies among the included studies, the data were analyzed using narrative synthesis, as the estimates of the studies are not directly comparable. Nonetheless, a quantitative summary of the accuracy across all included studies is provided to show overall trends.

### Risk of Bias

The risk of bias was assessed by two authors (MK & NvK) independently using the Quality Assessment of Diagnostic Accuracy Studies-2 tool (QUADAS-2)^26^. Any discrepancies were resolved through discussion again. QUADAS-2 uses four dimensions to rate the risk of bias and three dimensions to rate the applicability of a study to the research question. The risk of bias and applicability concerns were categorized into ‘low’, ‘some concerns’ and ‘high’.

## Results

### Included Studies

In total, 3019 potentially eligible studies were identified (3013 using the database search and 6 using citation search). After excluding ineligible studies, for example because they referred to emergency department triage only^27^, 19 studies were included in the review, see Figure 1.

**Figure 1.**
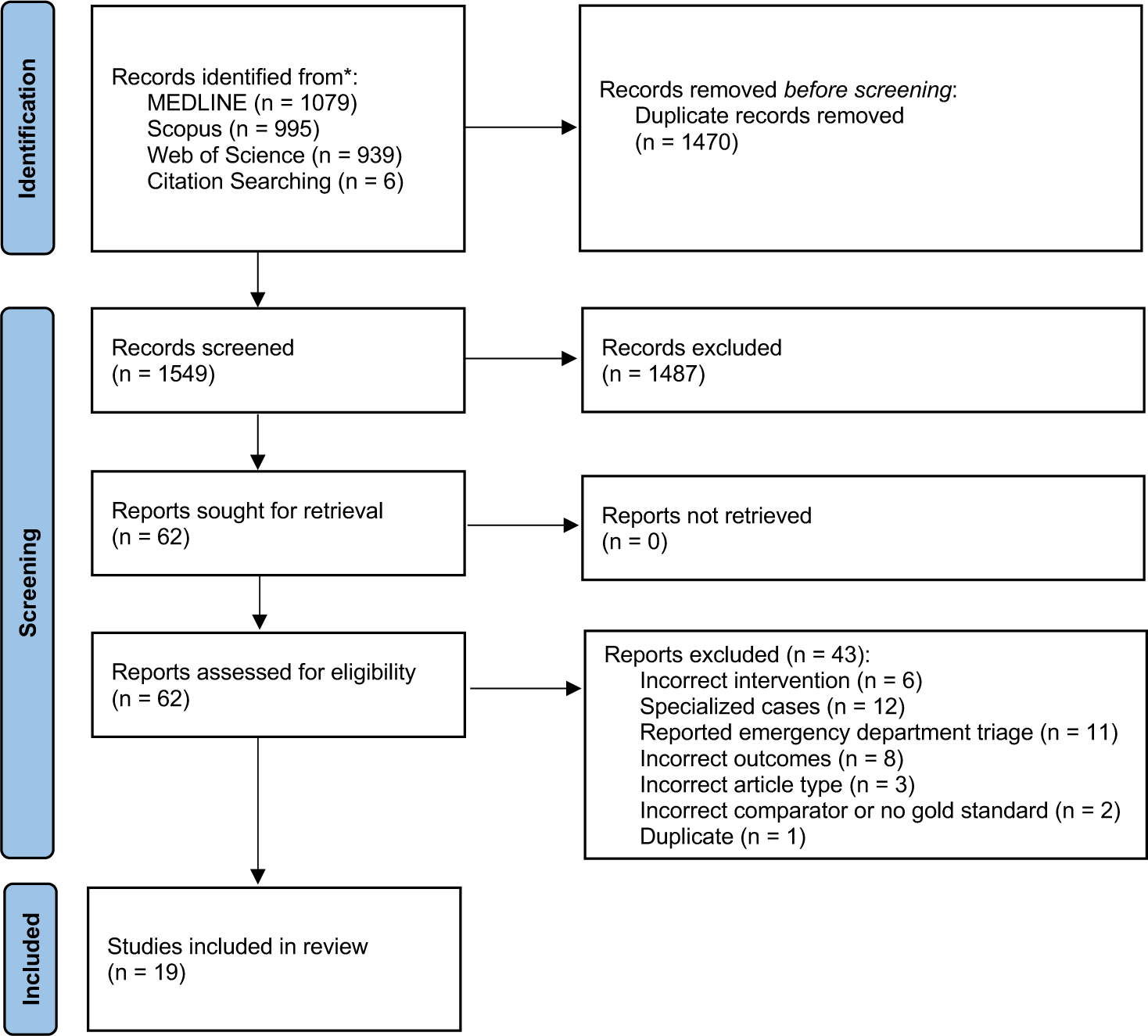
PRISMA flow diagram detailing the study search and selection process.

Most included studies (89%, 17/19) had at least one domain with a high risk of bias or some concerns, see Figure 2. The domain with the highest risk of bias was patient selection, as most studies used fictitious vignettes that were not based on real patient cases. For example, Semigran et al. used cases from text books and other medical resources that have a clear diagnosis assigned^2^ and other studies used cases that were completely made up by clinicians based on their experience^28–30^. Both methods do not represent real cases that SAAs are normally approached with^17,31,32^. Only five studies had a low risk of bias because they included real patient cases: three studies used patients from emergency departments and primary care settings^13,33,34^, one study directly surveyed SAA users^4^, and one study used real patient cases from medical laypeople that were actively making self-triage decisions and sought technical assistance for this decision^17^.

**Figure 2.**
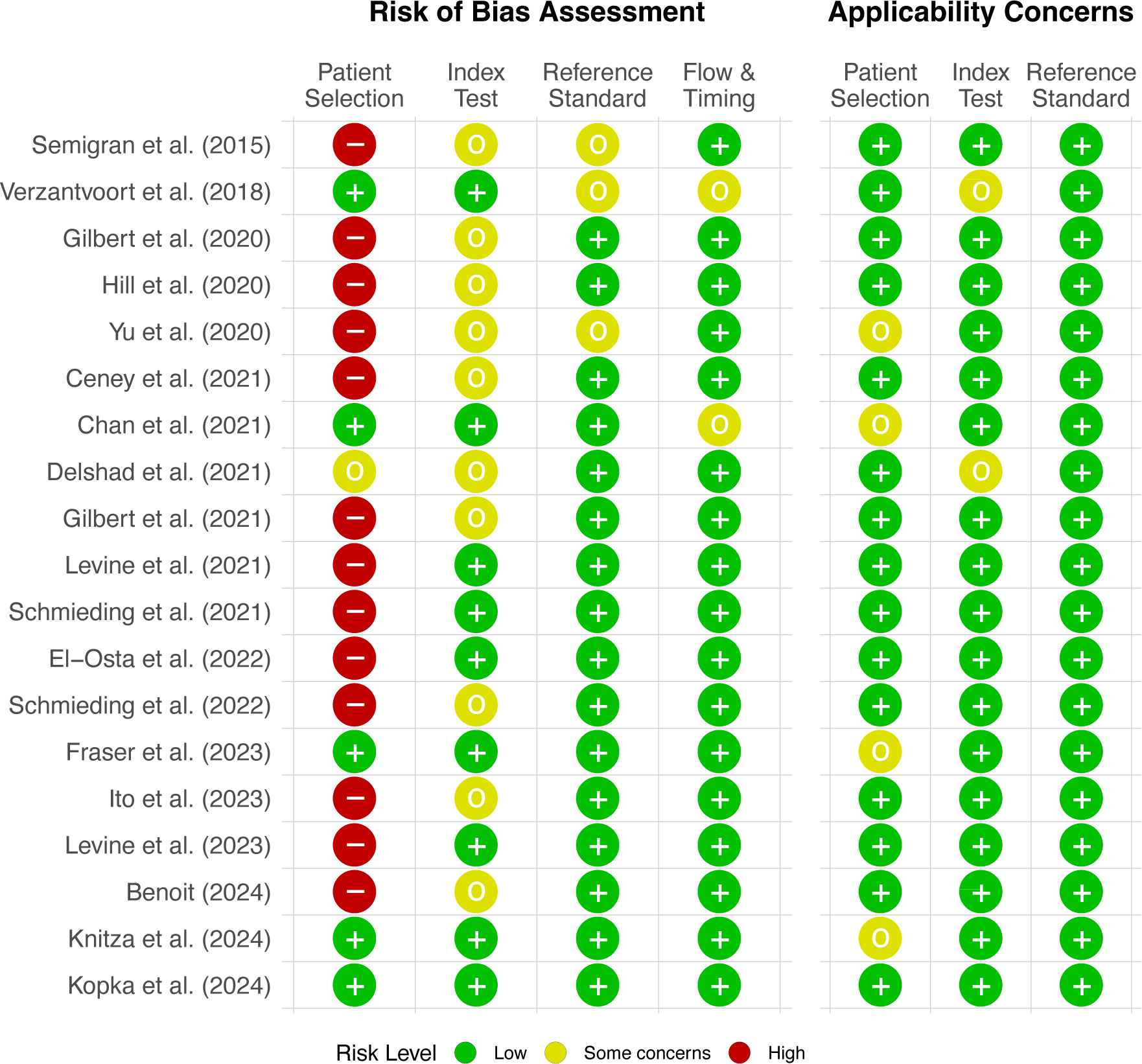
Risk of bias assessment and applicability concerns using QUADAS-2.

Index test was another domain in which many studies (53%, 10/19) have some risk of bias. Most of them did not report blinding of the inputter^2,12,21,22,29,35–39^. One study did not report how results from SAAs were obtained at all^36^. In the reference standard domain, only few studies have a moderate or high risk of bias (16%, 3/19). Those with concerns did not report how their gold standard was determined or used the judgement of one person only, e.g., the triage nurse in the emergency department^2,4,37^. Studies with some concerns regarding flow and timing had follow-up contact after several hours with a patient after using an app^4^ or did not mention when cases were reviewed^34^.

Applicability concerns were generally low. Most concerns comprised patient selection in studies that only used cases from the emergency department or a general practitioner setting, without including self-care cases^13,33,34,37^. Two studies had some concerns regarding the applicability of the index test, as one study used binary decisions only (visit a medical professional or not)^4^ and another study did not provide information how SAA results were determined^36^.

### Study Characteristics

In total, 14 (74%) studies analyzed the self-triage accuracy of SAAs^2,4,12,13,17,29,33–40^, four (21%) studies the accuracy of laypeople^17,23,30^, and four (21%) studies the accuracy of LLMs^17,21,22,28^. For SAAs, three (21%) studies let patients enter their symptoms directly^4,33,34^, three (21%) used real patient cases that were entered retrospectively^13,17,37^, and the remaining 8 (57%) studies used fictitious case vignettes developed by medical professionals^2,12,29,35,36,38–40^. For studies on laypeople, one study (25%) asked participants how they would rate the urgency of their own symptoms^34^, one (25%) used real patient cases that were presented to laypeople^17^ and two (50%) used fictitious vignettes phrased by medical professionals^23,30^. For LLMs, no study let patients enter symptoms themselves, one (25%) used real patient cases retrospectively^17^ and three (75%) used fictitious vignettes^21,22,28^.

Six (43%) studies examined only one SAA^4,33,34,36,39,40^, two (14%) studies examined two SAAs^13,37^ and six (43%) studies examined multiple SAAs^2,12,17,29,35,38^, ranging from seven^29^ to 23 different SAAs^2^. Studies on laypeople used sample sizes between 91 participants^23^ and 5000 participants^30^. For LLMs, three (60%) studies examined only one LLM^21,22,28^, whereas two (40%) studies examined multiple LLMs, ranging from two^13^ to five models^17^. The most frequently included SAA was Ada Health and the most frequently included LLM was GPT-4. All study characteristics are summarized in Table 1.

**Table 1.**
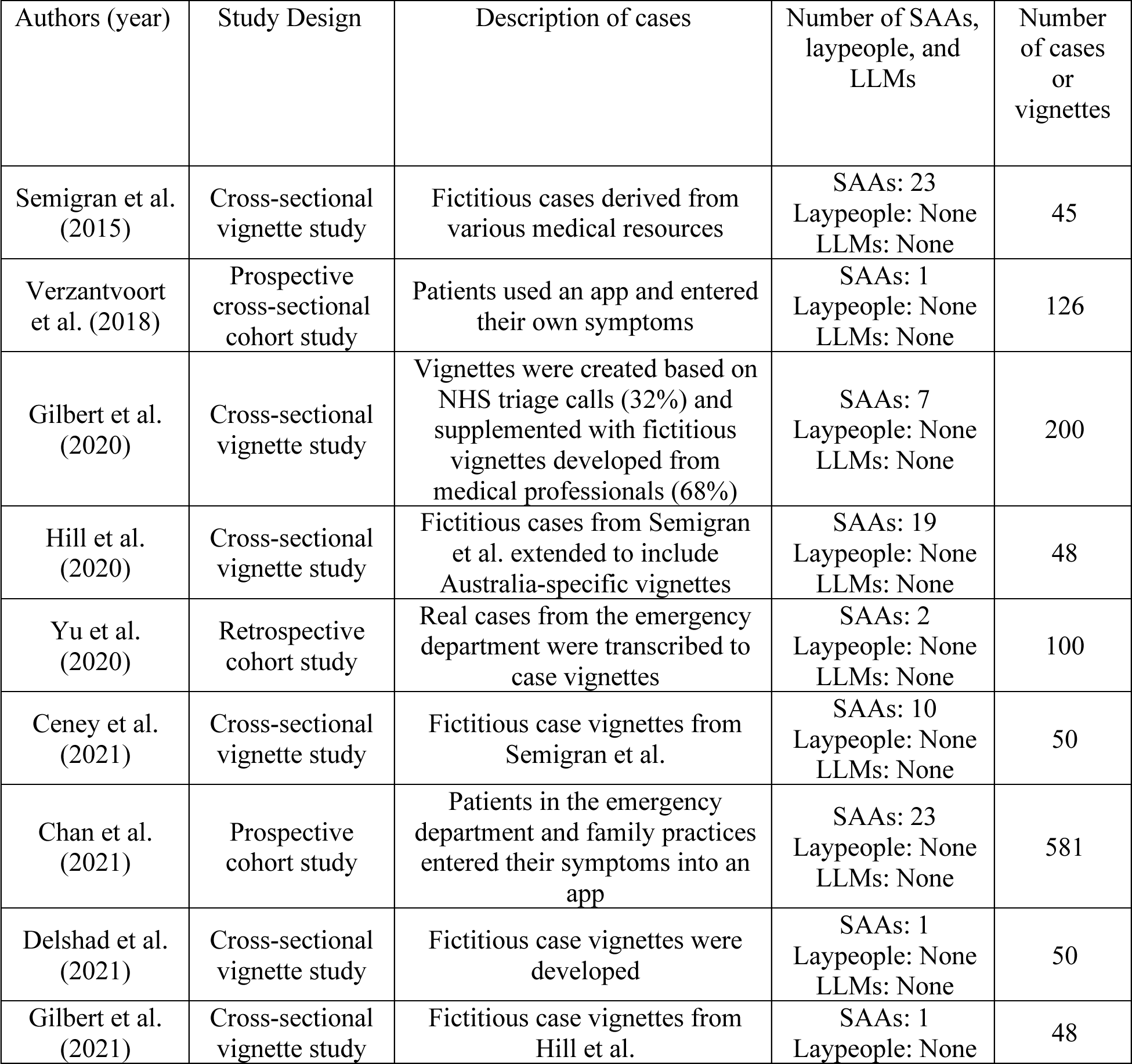
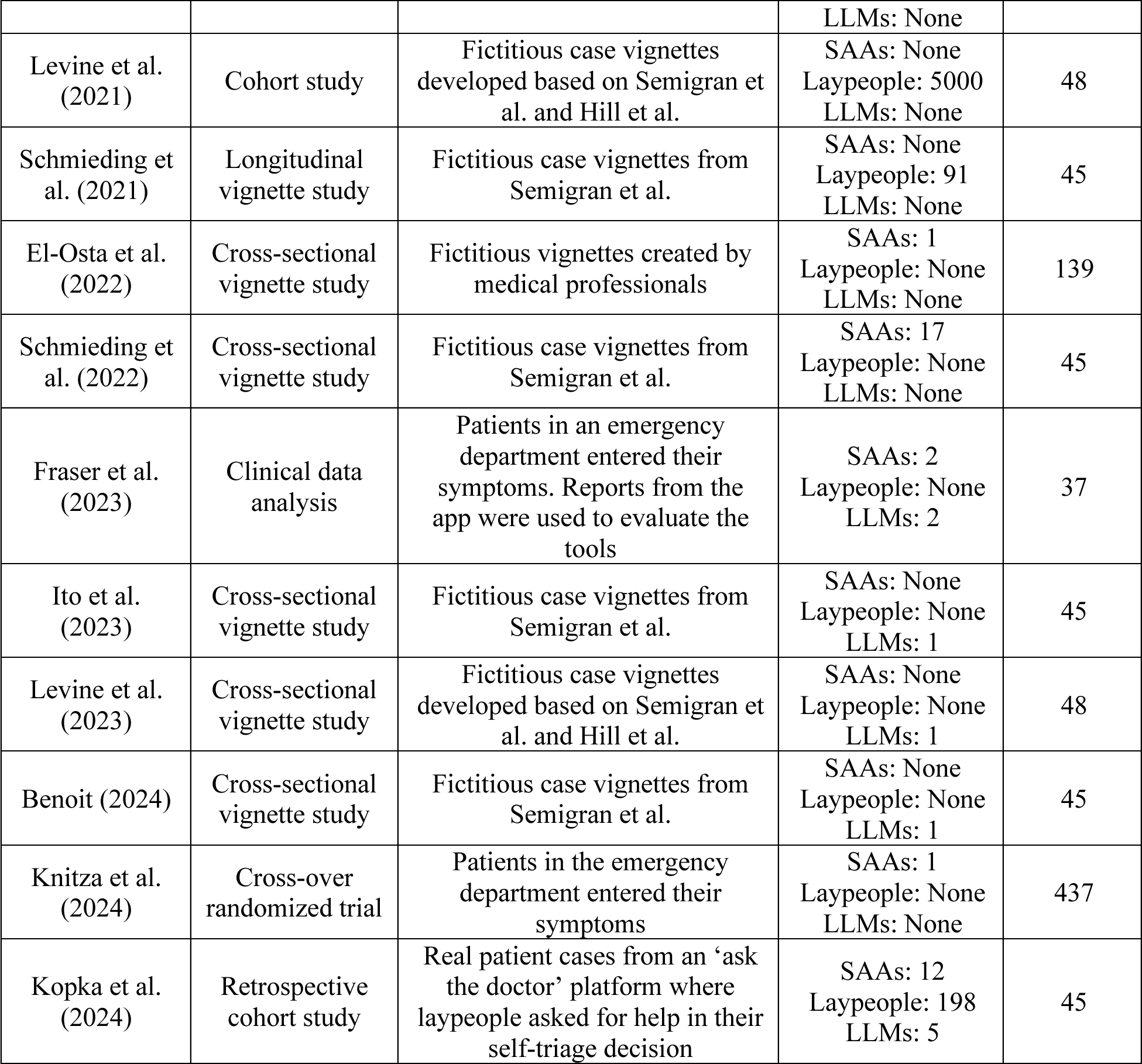
Characteristics of the included studies.

### Self-Triage Accuracy

The reported average accuracy of SAAs ranged from 25.9% in a study by Gilbert et al.^29^ to 88.0% in a study by Delshad et al.^36^, see Figure 3. However, the self-triage accuracy varies widely between different systems: The lowest individual SAA accuracy of 11.5% was reported in the study by Gilbert et al.^29^, while the highest accuracy of 90.0% was reported in a study by Ceney et al.^38^.

**Figure 3.**
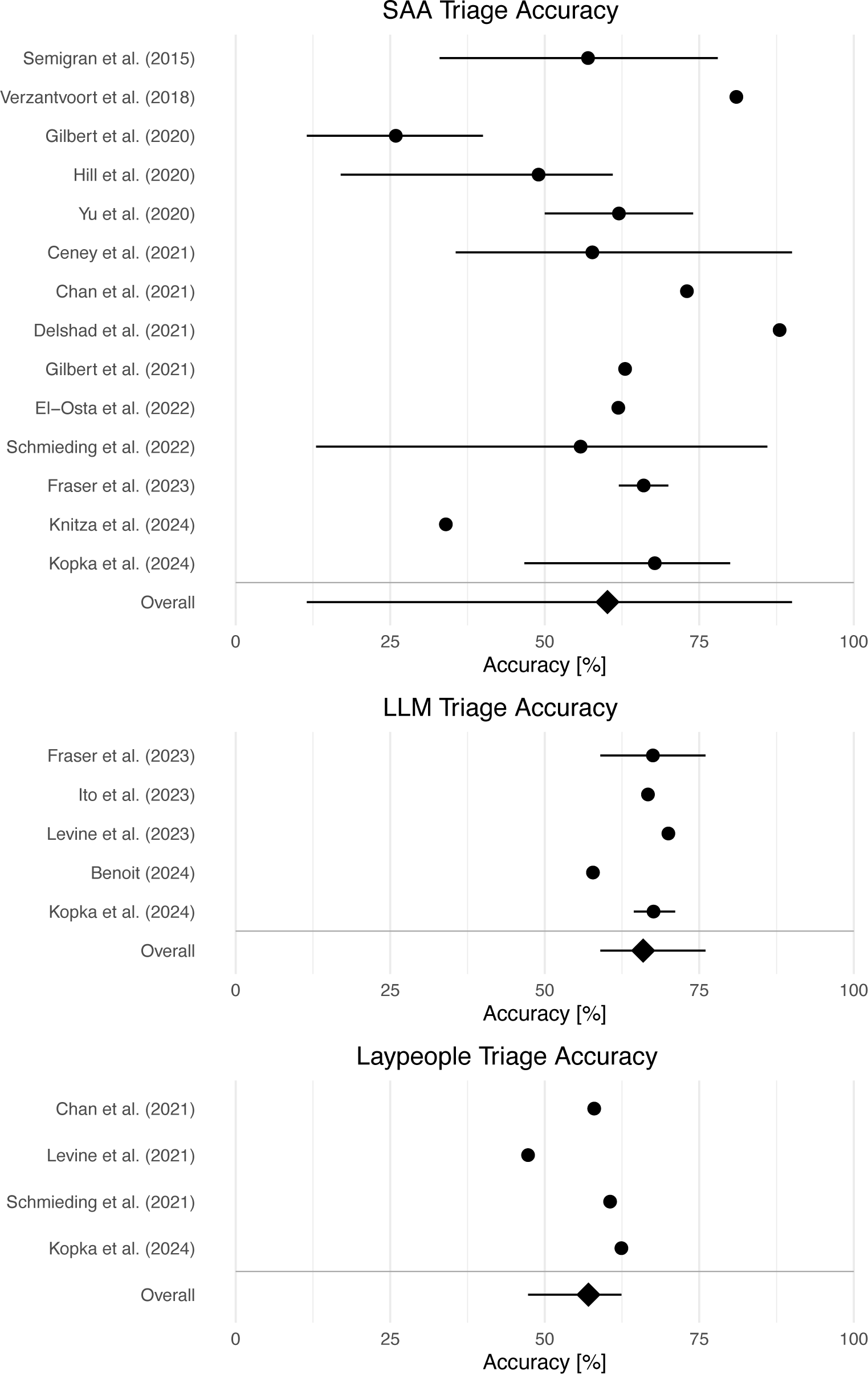
Overview of reported self-triage accuracy estimates for Symptom-Assessment Applications (SAAs), laypeople and Large Language Models (LLMs) *Note. Points indicate the reported mean and lines indicate reported minimum and maximum accuracy values within a study. Studies on laypeople reported means only without information on minimum and maximum values*.

The average accuracy of LLMs ranged from 57.8% in a study by Benoit^21^ to 70.0% in a study by Levine et al.^28^. Individual accuracy estimates for LLMs had a relatively low variation compared to SAAs and ranged from 57.8% in the study by Benoit^21^ to 76.0% in a study by Fraser et al.^13^.

The reported average accuracy of laypeople had a lower variation and ranged from 47.3% to 62.4%, see Figure 3. No study reported the individual accuracy, making a comparison of worst- and best-performing individuals with SAAs and LLMs impossible.

Most studies reported not only average accuracy but also average accuracy across different self-triage levels. For all three agents, accuracy differed between different urgency levels. SAAs generally had a high accuracy for emergency cases (74.5%, with a range from 57% to 100%) and a lower accuracy for urgent (53.3% range from 23.0% to 92.2%) and non-emergent cases (69.7%, range from 55.0 to 82.5%)^2,33,34,37^. Their accuracy was the lowest for self-care cases (42.1%, range from 0.0% to 74.0%)^4,33^.

LLMs had a moderate to high accuracy in emergency cases (66.7%, range from 50% to 86.7%) and reliably identified non-emergency cases (94.1%, range from 87% to 100%)^17,21,22,28^. However, they had a very low accuracy for self-care cases (10.8%, range from 6.15% to 16.7%)^17,28^.

Laypeople had a relatively high accuracy in identifying emergency cases (67.9%, range from 57.5% to 78.6%) and non-emergency cases (70.8%, range from 68.4% to 73.2%)^17,23,30^. For self-care cases, they had a low accuracy (35.6%, range from 25.4% to 46.7%)^23,30^, see Table 2.

**Table 2.**
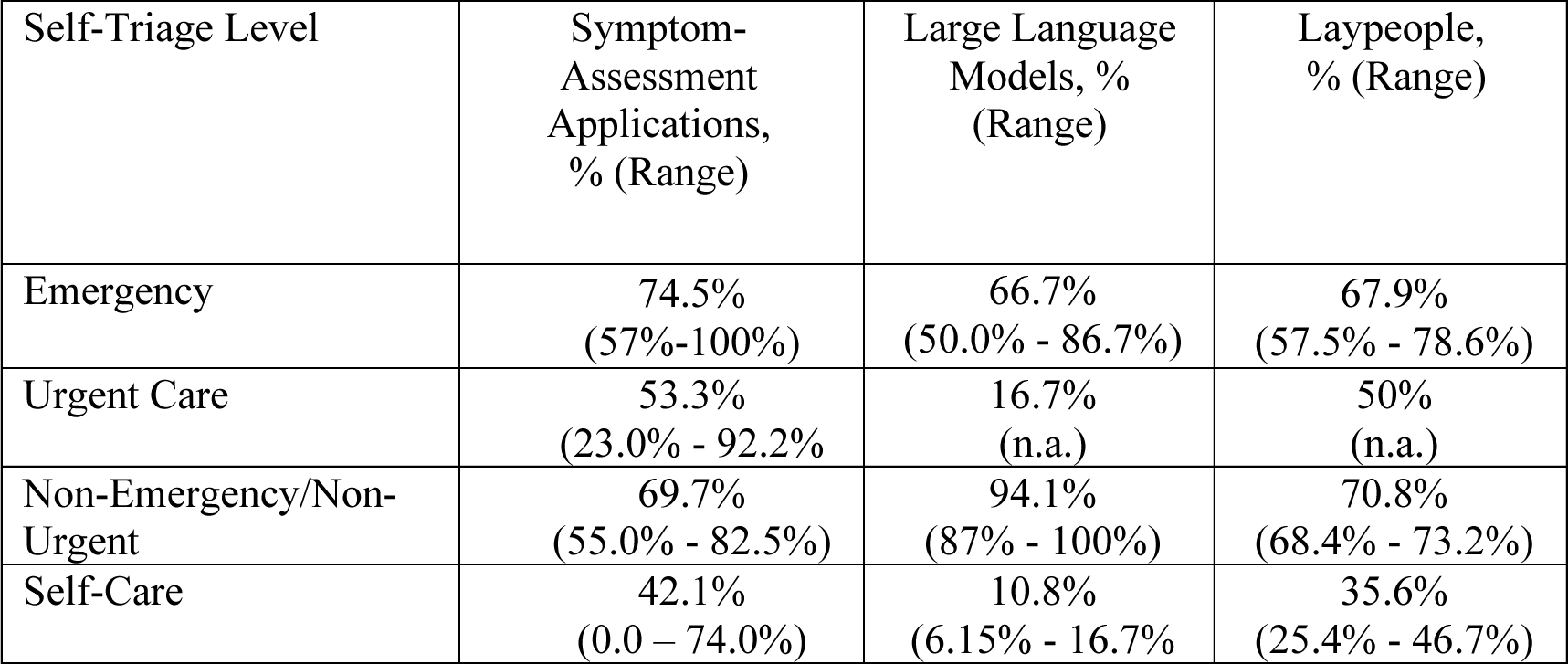
Reported self-triage accuracy of Symptom-Assessment Applications, Laypeople, and Large Language Models across different self-triage levels.

Individual SAAs demonstrated a high variability: Doctorlink – which was examined in one study only – had the highest accuracy with 90.0%, whereas K Health had the lowest accuracy with 21.5%. When only examining SAAs that were tested across multiple studies, Healthy Children (68.8%) and NHS111 online (66.1%) had the highest accuracy among all SAAs. The spread between the accuracy reported in different studies for the same SAA was high as well. For example, accuracy values for Symptomate ranged from 11.5% to 77.8% (with a mean of 48.6%), see Figure 4.

**Figure 4.**
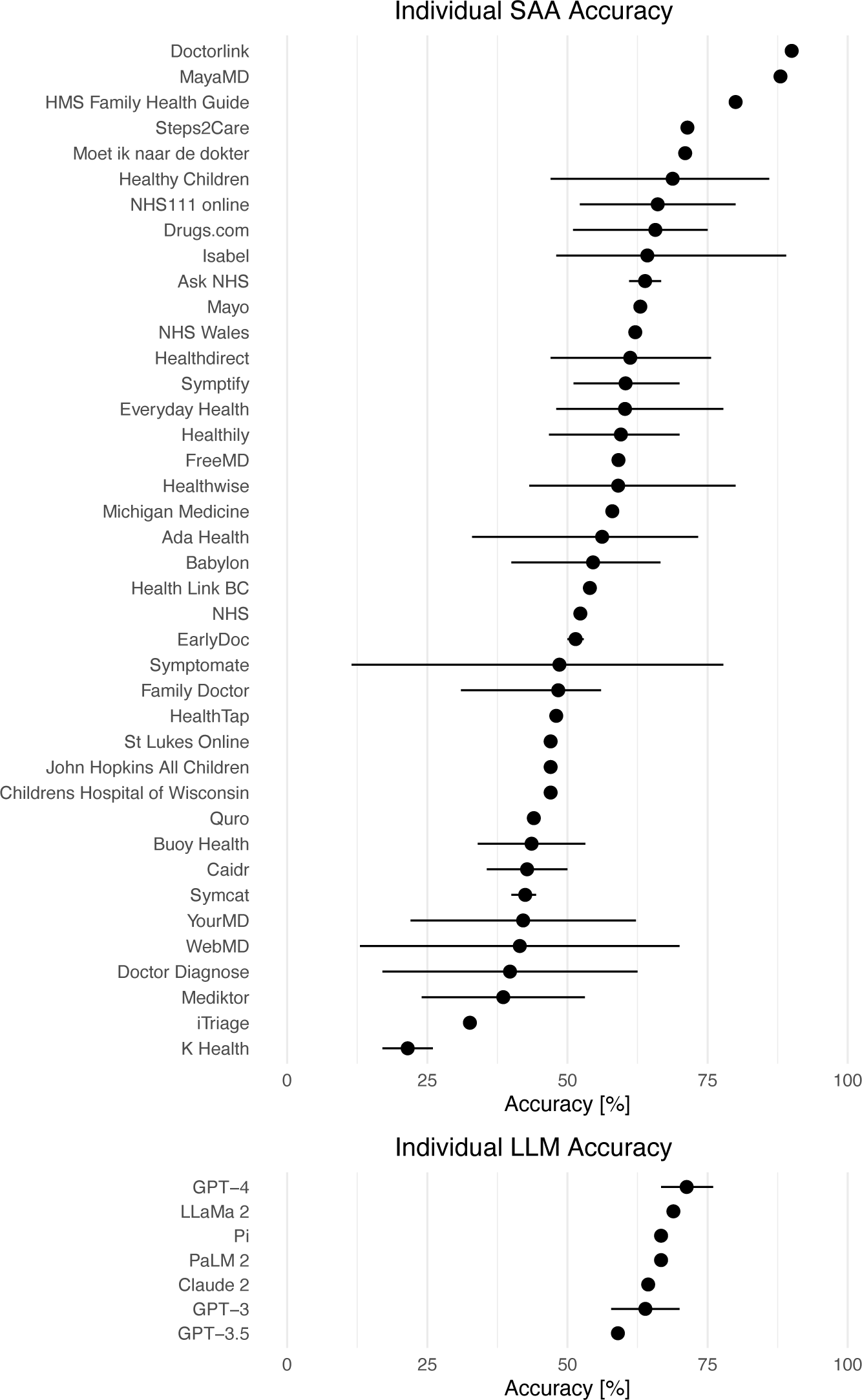
Overview of accuracy values reported for individual Symptom-Assessment Applications (SAAs) and large language models (LLMs) across multiple studies. *Note. Points indicate the mean of reported accuracy values and lines indicate minimum and maximum reported accuracy values. SAAs and LLMs without a line were examined in one study only. Since the methodology between studies differ and some are sponsored by the developer, the accuracy of these SAAs/LLMs should be interpreted with caution*.

For LLMs, the spread was relatively low. Although GPT-4 had the highest accuracy (71.3%), all of them scored between 59.0% and 71.3%. The accuracy between different studies only ranged from 66.7% to 76.0% for GPT-4, and 57.8% to 70.0% for GPT-3.

### Methodology

The methodology varied between studies. Although most studies assigned the gold standard for each case using a physician panel of two or more physicians that independently rated the cases and resolved disagreements through discussion^17,33,35^, some studies omitted independent ratings and directly used a physician discussion panel without letting them rate cases independently beforehand^29,36^. In other studies, the authors (who are physicians) assigned the gold standard themselves^28,30^ or used the decision of a single triage nurse^4,37^. Most studies used only one person to input data into SAAs and LLMs^2,12,13,21,22,28,35,38,39^. Two studies employed two people^17,37^, one study six people^40^ and one study eight people^29^. Some studies used medical professionals as inputters^2,29^, while others specifically let laypeople enter the symptoms^12,17^. Notably, only two studies mentioned blinding inputters to the gold standard solution^13,17^. Although most studies used three self-triage levels in their assignment^2,12,13,17,21–23^, some used only two^4,37^ (e.g., emergency or no emergency), and one study even used six levels^29^. Additionally reported self-triage outcomes varied between studies: One study used metrics from signal detection theory^4^, three studies reported the comprehensiveness of an SAA^17,29,38^, seven studies reported the inclination to over-/ and undertriage^12,13,17,23,33,34,37^, and five studies reported the safety of advice^13,17,29,38,40^. One study additionally reported the Capability Comparison Score, which was developed specifically to compare SAAs^17,41^.

**Table 1.** Methodological details of the included studies.

| Authors (year) | Gold Standard | Number of inputters | Number of self-triage levels | Other outcomes reported |
| --- | --- | --- | --- | --- |
| Semigran et al. (2015) | Correct diagnosis was part of medical resource; no information on self-triage level | 1 | 3 | None |
| Verzantvoort et al. (2018) | Triage nurse determined self-triage level after telephone interview | n.a. | 2 | Sensitivity, Specificity, Positive Predictive Value, Negative Predictive Value |
| Gilbert et al. (2020) | Assigned by physician panel | 8 | 6 | Comprehensiveness, Safety |
| Hill et al. (2020) | Two physicians and one emergency specialist rated cases, disagreement resolved through discussion | 1 | 4 | None |
| Yu et al. (2020) | Assigned triage level by triage nurse upon visiting emergency department | 2 | 2 | Over-/Undertriage |
| Ceney et al. (2021) | Taken from Semigran et al. and assessed against National Institute for Health and Care Excellence summaries | 1 | 4 | Comprehensiveness, Safety |
| Chan et al. (2021) | Decision from treating physician was reviewed by two physicians | n.a. | 4 | Over-/Undertriage |
| Delshad et al. (2021) | Several physicians from different institutions were asked about the most appropriate self-triage level. They were asked to develop a consensus | n.a. | 4 | None |
| Gilbert et al. (2021) | Taken from Hill et al. | 1 | 4 | None |
| Levine et al. (2021) | Assigned by two physicians | n.a. | 4 | None |
| Schmieding et al. (2021) | Taken from Semigran et al. | n.a. | 3 | Over-/Undertriage |
| El-Osta et al. (2022) | Multiple gold standards tested: general practitioners (GPs) that developed vignettes also assigned solution. 3 independent GPs were asked about correct self-triage level. Both solutions were pooled | 6 | 3 | Safety |
| Schmieding et al. (2022) | Taken from Semigran et al. | 1 | 3 | Over-/Undertriage, Binary Self-Triage decision |
| Fraser et al. (2023) | Three emergency department physicians rated each case | 1 | 3 | Safety, Overcaution |
| Ito et al. (2023) | Taken from Semigran et al. | 1 | 3 | None |
| Levine et al. (2023) | Assigned by two physicians | 1 | 4 | None |
| Benoit (2024) | Taken from Semigran et al. | 1 | 3 | None |
| Knitza et al. (2024) | Two physicians rated each case | n.a. | 4 | Over-/Undertriage |
| Kopka et al. (2024) | Panel of 2 physicians rated independently, disagreement resolved through discussion | 2 | 3 | Safety, Over-/Undertriage, Comprehensiveness, Capability Comparison Score |

## Discussion

This systematic review aimed to synthesize available evidence on self-triage accuracy, focusing not only on SAAs but also on LLMs as an alternative, and on laypeople as the user group. Our findings indicate that SAAs have a relatively low accuracy on average, but they also show that accuracy is highly dependent on the specific tool used. Most studies report a high spread between different SAAs and there is also high heterogeneity between the studies. However, when assessing individual SAAs across different studies, some tools seem to consistently perform well. For example, NHS 111 online was included in multiple studies and consistently showed moderate to high accuracy. Conversely, Mediktor showed a consistently low performance across multiple studies. Surprisingly, LLM accuracy does not have a high spread in comparison – all studies report values between 58% and 76% and the individual spread for LLMs across studies is minimal as well. The same holds true for laypeople: All the included studies report an accuracy between 47% and 62%, indicating that laypeople make decisions better than chance level, but far from perfect.

Our review, while including more recent studies, aligns with the findings from previous systematic reviews. These reviews consistently report that SAA accuracy is relatively low, but note that the variability between the tools is very high^14–16^. This variation is understandable, considering that SAAs are developed by different institutions, each using different methods and working with varying levels of funding. For example, some developers use simple rule-based algorithms, while others use Bayesian networks^42^. Based on varying accuracy levels, all reviews conclude that SAAs pose a safety risk and suggest that their use might not be encouraged. While the safety concerns are valid and important, it is noteworthy that laypeople also tend to make decisions with only moderate accuracy. Hence, leaving them unassisted in their self-triage decisions might not be a viable solution either and the interaction between laypeople and digital tools for self-triage warrants further investigation. Because the interaction is not fully understood, there’s a risk that their combined errors could lead to even worse decisions than if each made a decision separately. Alternatively, their correct decisions might complement each other and increase the overall self-triage accuracy beyond the accuracy of each agent alone. Since humans make the final decision in the end, it is also important to understand how they include and compensate incorrect advice. One previous study on human-SAA interaction suggests that laypeople can increase their accuracy with well-performing SAAs, but not to the level of the SAA’s isolated accuracy^43^. However, users were able to compensate incorrect recommendations and were not entirely dependent on the system. Thus, the study overall suggests that errors do not add up, but rather that laypeople can successfully use SAAs – even if the system’s accuracy is not perfect – and compensate incorrect recommendations.

When comparing SAAs, LLMs and laypeople, it is also important to examine the specific decisions that are made. The accuracy of all three agents differed drastically between the urgency levels of the presented cases. Whereas all performed relatively well in identify emergencies (with laypeople and SAAs showing very similar accuracy), their accuracy in self-care cases varied drastically. SAAs had a variation between 0 and 74%, while laypeople solved between 25% and 47% of these cases correctly. LLMs rarely advised self-care at all and thus had an accuracy below 20%. These findings indicate that laypeople may not require assistance in identifying emergencies but could profit from support in identifying self-care cases. However, LLMs are not well-suited for this task and only certain SAAs can be helpful in this regard. Some previous studies suggest dividing the urgency levels into two steps to better reflect how laypeople make self-triage decisions: First, they determine whether their symptoms require medical attention at all, and if so, they then decide where to seek care^12,44^. Considering our findings, laypeople may need more assistance in determining whether their symptoms require medical attention rather than deciding where to seek care; this could be the decision in which SAAs and other tools could be more beneficial. Thus, it is not universally advisable to recommend or dismiss using SAAs. Rather, recommendations should depend on the specific implementation use case. When deciding between emergency and non-emergency care, LLMs might be helpful due to their high accuracy in this regard. However, when deciding if care is needed at all, LLMs generally do not offer any assistance and only some SAAs are useful.

For users, a general recommendation for using any SAA or LLM is not advisable. However, some tools might be helpful depending on the specific decision they need to make. For instance, when deciding between emergency and non-emergency care, LLMs and specifically GPT-4 might be beneficial, as it has been found to be relatively safe and accurate in this decision^13,17^. On the other hand, if users want to determine whether their symptoms warrant any medical attention at all, using a tool like NHS 111 online could be helpful due to its high accuracy in this decision. Nevertheless, users should always use these tools with caution and cross-verify the recommendations with additional information sources and critical thinking.

For evaluators (such as other researchers, implementers or policymakers), a standardized evaluation process is essential. The primary quality risk in current evaluations is the use of fictitious case vignettes that do not represent real patients^15–17,45^. Although using these vignettes is convenient and resource-efficient, they often yield results that are not generalizable to real-world settings^17,45^. Since using real patients who enter their own symptoms might not be feasible and cannot be applied to evaluate multiple SAAs, a cost-effective alternative could involve using real patient descriptions that are entered into SAAs. A procedure for that is available with the RepVig framework^17^. Afterwards, specific SAAs can be tested with actual patients in a clinical trial to validate positive findings. Alongside the type of cases used in testing SAAs, other methodological variations influence the outcomes, such as the number of inputters, the gold standard assignment, the metrics, and the number of self-triage levels that are reported. Although recent studies provide specific recommendations for these issues^17,40,41,46^, they are rarely being applied yet. For example, Meczner et al. examined inputter variability and suggest using standardized instructions, multiple inputters, and a pooled accuracy metric to reflect the recommendations that multiple inputters receive^46^. El-Osta et al. investigated the gold standard assignment process and concluded that pooling decisions of two independent physician panels gets closer to the best solution than using one physician panel or a single person only^40^. Kopka et al. reviewed the metrics reported in other studies and proposed a set of standardized metrics to better understand the strengths and weaknesses of an SAA^41,45^. Lastly, standardizing the number of self-triage levels could improve comparability both within and between studies. Most studies use three or four levels, yet not all SAAs provide an ‘urgent care’ recommendation^12^. Thus, we suggest that using three triage levels – as originally proposed by Semigran et al.^2^ – might increase comparability.

This review has several limitations. First, unlike previous systematic reviews, we focused solely on self-triage accuracy rather than diagnostic accuracy. This choice was motivated by the relevance to laypeople: While a preliminary diagnosis might lead to further information-seeking, a correct diagnosis often requires medical tests or more details that are not accessible to laypeople^47^. Ultimately, diagnoses are made by medical professionals anyway. As noted in several studies already, aiding laypeople in finding the most suitable care pathway is a more effective use case for these tools^2,15^. This perspective is also reflected in the included studies, as only one study involving laypeople assessed their diagnostic accuracy^30^ – unliked numerous SAA studies that typically evaluate both diagnostic and self-triage accuracy^15,16^.

Another limitation concerns the number of included studies: Although many studies test the accuracy of SAAs, only few studies examine the accuracy of laypeople. A potential reason might be the novelty of the field, and that researchers thus initially focus on evaluating the technological aspects before progressing to more realistic scenarios that include human participants – akin to lab studies that first assess effects under controlled conditions and then move on to observational studies to confirm these effects in the real world. Similarly, the number of studies evaluating LLMs was also low. Because LLMs were first released to the public with ChatGPT in 2022, the technology can be considered relatively new and there has been limited time to conduct and publish studies on their accuracy. Although these is a vast body of medical research on LLMs already, most of it has focused on their ability to pass pre-specified exams like board tests or other diagnostic tasks^20,48^. As more time passes, we can expect to see additional evidence on the self-triage accuracy of LLMs and conducting an updated systematic review on their accuracy might be insightful. This is particularly relevant, because LLMs seem to quickly improve their accuracy across various tasks with new iterations^49^.

Lastly, the methodologies varied among the included studies, which makes the direct comparison of accuracy estimates complicated. While differences in methods are more pronounced for diagnostic accuracy – e.g., some studies evaluate only the first diagnosis while others consider the top 3, 5, or 10^15,16^ – the methods for self-triage accuracy also vary. A major issue concerns using fictitious vignettes in most studies that were phrased by clinicians and developed based on clear case descriptions from medical education resources or from physicians’ experience. Although these vignettes represent clear cases with a definitive solution, they do not accurately reflect real cases that SAAs are approached with^17,32,50,51^. As a result, generalizability of most included studies is questionable.

## Conclusions

In conclusion, the performance of SAAs compared to laypeople varies; some SAAs outperform laypeople, while others do not. Therefore, universally recommending SAAs to the public may not be advisable, but well-performing SAAs might warrant a recommendation if their safety is assured. LLMs showed less variability and higher accuracy than many SAAs in handling both emergency and non-emergency cases, which suggests a potential usefulness in these scenarios. Nonetheless, they rarely recommend self-care and can thus not be universally endorsed either.

Deciding which tools to use should be based on the specific use case. For users confident that their symptoms require medical attention, a high-performing SAA or LLM could be beneficial. However, for those uncertain whether their symptoms warrant medical attention at all, SAAs that effectively differentiate between self-care and medical care could be useful, while LLMs in their current form do not provide any assistance in this decision-making process. Although general endorsement of SAAs or LLMs is not recommended, their use should not be outright discouraged either. The appropriateness of these tools depends on the specific use case and the particular tool that is considered.

## Author contributions

MK conceived of the study. MK and NvK conducted the screening and data extraction. MK conducted the data analysis and wrote the first draft of the manuscript. All authors provided critical input and worked on manuscript development.

## Data availability

The search strategy can be found in the Methods section and all information on studies are cited. Any additional data is available upon request.

